# Evaluating the Long-Term Efficacy of COVID-19 Vaccines

**DOI:** 10.1101/2021.01.13.21249779

**Authors:** Dan-Yu Lin, Donglin Zeng, Peter B. Gilbert

**Author notes:** Correspondence: Dan-Yu Lin, PhD, Department of Biostatistics, University of North Carolina, Chapel Hill, NC 27599-7420, USA.

## Abstract

Large-scale deployment of safe and durably effective vaccines can curtail the COVID-19 pandemic.^1−3^ However, the high vaccine efficacy (VE) reported by ongoing phase 3 placebo-controlled clinical trials is based on a median follow-up time of only about two months^4−5^ and thus does not pertain to long-term efficacy. To evaluate the duration of protection while allowing trial participants timely access to efficacious vaccine, investigators can sequentially cross participants over from the placebo arm to the vaccine arm according to priority groups. Here, we show how to estimate potentially time-varying placebo-controlled VE in this type of staggered vaccination of participants. In addition, we compare the performance of blinded and unblinded crossover designs in estimating long-term VE.

**Authors’ Information:** Dan-Yu Lin, Ph.D., is Dennis Gillings Distinguished Professor of Biostatistics, and Donglin Zeng, Ph.D., is Professor of Biostatistics, Gillings School of Global Public Health, University of North Carolina, Chapel Hill, NC 27599-7420, USA. Peter B. Gilbert, Ph.D., is Member, Vaccine and Infectious Disease Division, Fred Hutch, Seattle, WA 98109-1024, USA.

**Summary:** We show how to estimate the potentially waning long-term efficacy of COVID-19 vaccines using data from randomized, placebo-controlled clinical trials with staggered enrollment of participants and sequential crossover of placebo recipients.

## Introduction

A number of studies have been conducted around the world to evaluate the efficacy and safety of investigational vaccines against novel coronavirus disease-2019 (COVID-19).^4−12^ Interim results from several large-scale phase 3 randomized, observer-blinded, placebo-controlled clinical trials have demonstrated high VE,^4−6^ far exceeding the FDA and WHO thresholds of 50% reduction of symptomatic disease.^13−14^ However, those trials have been underway for only several months, and the data they have collected thus far can only speak to short-term VE. For example, the recently published results for the Pfizer/BioNTech vaccine BNT162b2 and the Moderna vaccine mRNA-1273 are based on a median follow-up time of approximately two months after the second dose;^4−5^ therefore, the reported 94-95% VE pertains only to an average of two months post vaccination.

Vaccine effect can wane over time because of declining immunologic memory or changing antigenicity of the pathogen. A vaccination can be followed with booster doses to maintain a protective level of immunity among susceptible individuals, but the nature of the protection over time must be understood so that an effective vaccination and boosting schedule can be determined. Thus, after FDA issues an Emergency Use Authorization (EUA), vaccine sponsors should continue to collect placebo-controlled data on primary endpoints in any ongoing trials for as long as feasible.^15^

Although continuing blinded follow-up of the original treatment arms is the ideal way to evaluate long-term efficacy and safety, placebo recipients should be offered the vaccine at some point after an EUA. One strategy is the “rolling crossover”, which vaccinates placebo recipients around the same time as general population members in the same priority tier. Under this design, placebo participants are vaccinated at different times, with the timing of vaccination depending on enrollment characteristics that define their priority tier.

Because participants are vaccinated at different times and community transmission of SARS-CoV-2 virus changes over time, existing statistical methods — which assume that trial participants are vaccinated at the same time or that community transmission is constant over time — do not produce valid estimates of long-term VE in the presence of waning vaccine effect. In fact, participants in COVID-19 vaccine trials are enrolled over several months and thus are randomized to the vaccine and placebo groups at different times; therefore, the timing of vaccination varies among vaccinees even if the placebo group is maintained throughout the study.

At the time of interim analysis, there were 8 and 162 symptomatic COVID-19 cases in the BNT162b2 vaccine and placebo groups, respectively,^4^ such that standard methods would estimate VE at 95%. Because of staggered enrollment and time-varying community transmission, this estimate is difficult to interpret if the VE wanes over time. (Adjustment for the participant’s follow-up time would not change the estimate since the person years of follow-up are the same between the vaccine and placebo groups. Due to relatively low disease incidence, standard Poisson and Cox models would yield the same estimate.^16^)

In this article, we show how to properly assess the durability of VE under staggered enrollment and time-varying community transmission, allowing higher-risk placebo volunteers to get vaccinated earlier than lower-risk ones during the crossover period. Our framework provides unbiased estimation of the entire curve of placebo-controlled VE as a function of time elapsed since vaccination, up to the point where most of the placebo volunteers have been vaccinated. We investigate the bias and precision of our approach in estimating long-term VE under various crossover designs, including blinded crossover, in which participants do not know the order of treatments they receive, and unblinded crossover, in which participants are notified of their randomization assignments at the time of crossover. We also discuss how to perform sensitivity analysis when unblinded follow-up data are used.

## Methods

Figure 1 provides a schematic illustration of the rolling crossover strategy in the context of COVID-19 vaccine trials. In this scenario, participants are screened and randomly assigned to vaccine or placebo over a 4-month period, the vaccine is granted EUA in the 5th month on the basis of interim results, and crossover occurs over the next five months. This design provides information about VE for 10 months. We aim to estimate time-varying VE under any crossover design, up to the point when there are very few placebo participants left.

**Figure 1.**
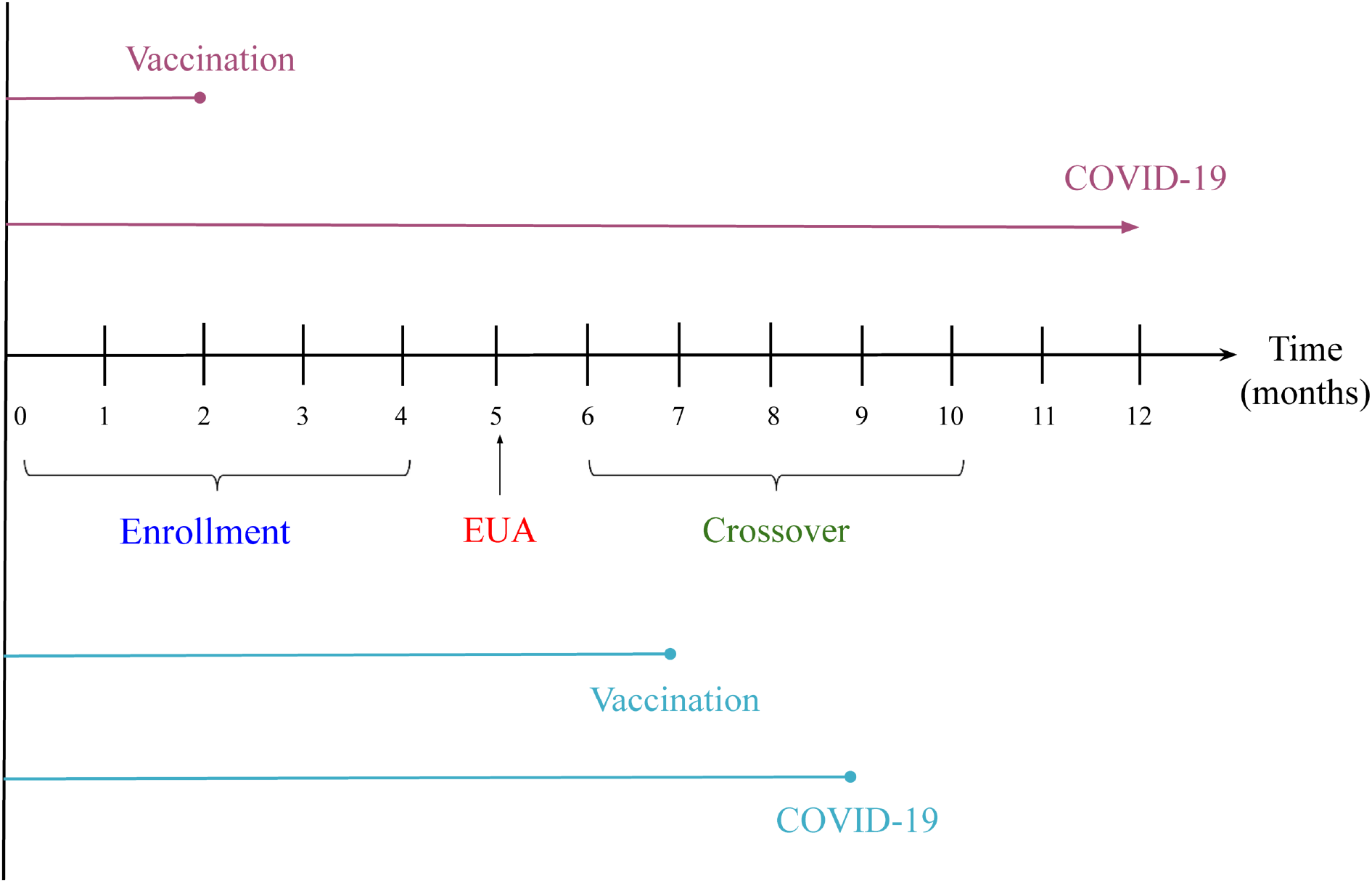
A phase 3 COVID-19 vaccine trial. Participants are enrolled over a 4-month period, EUA is issued at month 5, and crossover occurs over months 6–10. The top two lines represent a participant who is vaccinated at month 2 and does not develop symptomatic COVID-19 during the trial, and the bottom two lines represent a participant who is vaccinated at month 7 and develops symptomatic COVID-19 at month 9.

The endpoint of interest is time to symptomatic COVID-19 disease. We allow the risk of disease to vary over the calendar time and to depend on baseline risk factors, such as age, sex, ethnicity, race, occupation, and underlying health conditions; we allow the effect of vaccine on disease occurrence to depend on the time elapsed since vaccination.

We consider two definitions of time-varying VE: (1) day-*t* VE is the percentage reduction in the hazard rate or instantaneous risk of disease at day *t* for those who were vaccinated *t* days ago compared with those who have not been vaccinated; and (2) *t*-day VE is the percentage reduction in the attack rate or cumulative incidence of disease over the *t*-day period for those who were vaccinated at the start of the period compared with those who were unvaccinated throughout the period. We denote these two VE measures by VE_h_ and VE_a_, where h and a stand for hazard rate and attack rate, respectively. These two definitions are equivalent when the effect of vaccine is constant over time. If the effect of vaccine wanes over time, then VE_a_ is larger than VE_h_.

In Supplemental Appendix 1, we formulate the above concepts through an adaptation of the well-known Cox regression model,^17−18^ in which each participant’s time to disease occurrence is measured from a common origin, namely the start of the clinical trial, and the hazard ratio of vaccine versus placebo depends on the time elapsed since vaccination. We formally define VE_a_ as one minus the time-averaged hazard ratio, which is approximately the ratio of the cumulative incidence. We derive the maximum likelihood estimator for VE_a_ as a function of time elapsed since vaccination. We show that the estimator is approximately unbiased and normally distributed, with a variance that can be estimated analytically, enabling one to construct valid confidence intervals for the VE_a_ curve. Finally, we propose a method to estimate VE_h_ by kernel-smoothing the estimate of VE_a_.

## Results

We conducted a set of simulation studies mimicking the BNT162b2 vaccine trial. We considered 40,000 participants, who entered the trial at a constant rate over a 4-month period and were randomly assigned to vaccine or placebo in a 1:1 ratio. The vaccine received an EUA from FDA at the 5th month, by which time there were about 300 COVID-19 cases in the placebo group. To reflect the increase of COVID-19 cases since last summer and the expected downward trend in the spring due to vaccine rollout and other factors, we let the disease risk increase over the first 7 months and decrease afterward. We chose three combinations of 5-month VE and 10-month VE: (a) VE_a_(5 mos.) = VE_a_(10 mos.) = 95%; (b) VE_a_(5 mos.) = 85%, VE_a_(10 mos.) = 75%; and (c) VE_a_(5 mos.) = 70%, VE_a_(10 mos.) = 50%.

We considered the statistically optimal design of keeping all participants on their original treatment assignments until the end of the trial. We refer to this design as Plan A and regard it as a benchmark. We also considered three blinded crossover designs:

B. Crossover starts at month 6, 7, 8, 9, or 10 for participants with priority tier of 1, 2, 3, 4, or 5, respectively, with each participant’s waiting time for the clinic visit following the exponential distribution with mean of 0.5 month.

C. 20% of participants follow Plan A, and the rest follow Plan B.

D. Crossover starts at month 6 for all participants, with the waiting time following the exponential distribution with mean of 0.5 month.

Both B and C are priority tier-dependent rolling crossover designs. The difference is that under Plan B, all placebo recipients cross over to the vaccine arm, whereas under Plan C, 20% of participants choose for altruistic reasons to stay on their original treatment assignments. Under Plan D, all placebo recipients are vaccinated quickly without any priority tiering. With blinded crossover, placebo participants receive the vaccine and vaccine participants receive the placebo at the point of crossover; none of the participants are aware of the order of their treatments. All participants are followed until the end of the trial or the time of analysis, which is 10.5 months since trial initiation. The designs of these simulation studies are detailed in Supplemental Appendix 2.

The results for the estimation of VE_a_ based on 10,000 simulated datasets are summarized in Table 1 and Figure 2. The proposed method yields virtually unbiased estimates of the VE_a_ curves over the 10-month period for Plans A–C in all three scenarios of long-term VE; it also yields accurate variance estimates, such that the confidence intervals have correct coverage probabilities. When VE is constant over time, the standard errors for the estimates of VE_a_ under Plans B and C are slightly lower than those of Plan A. When VE wanes over time, the standard errors for the estimates of 5-month VE_a_ under Plans B and C are also slightly lower than those of Plan A; however, the standard errors for the estimates of 10-month VE_a_ under Plans B and C are higher than those of Plan A, with the standard errors being slightly lower under Plan C than under Plan B. Under Plan D, the estimates of 10-month VE_a_ may be slightly biased, with higher standard errors than under Plans A–C; these results are not surprising, because under this plan, the number of unvaccinated participants diminishes rapidly after month 6.

**Table 1.** **Estimation of Time-Varying** VE_a_ **by the Proposed and Standard Methods Under No Crossover (A) and Three Blinded Crossover Plans (B–D)**

| Scenario | Plan | Time | True | Proposed method |  |  |  | Standard |  |
| --- | --- | --- | --- | --- | --- | --- | --- | --- | --- |
| | | | $VE_a$ | Mean | SE | SEE | CP | Mean | SE |
| (a) 5-mos $VE_a = 95\%$<br>10-mos $VE_a = 95\%$ | A | 5 mos | 95% | 95.0% | 0.8% | 0.8% | 95.2% | | |
|  |  | 10 mos | 95% | 95.0% | 0.8% | 0.8% | 94.5% | 95.0% | 0.6% |
|  | B | 5 mos | 95% | 95.0% | 0.7% | 0.7% | 95.1% |  |  |
|  |  | 10 mos | 95% | 95.1% | 0.8% | 0.7% | 94.5% | 95.0% | 0.6% |
|  | C | 5 mos | 95% | 95.0% | 0.7% | 0.7% | 95.2% |  |  |
|  |  | 10 mos | 95% | 95.0% | 0.7% | 0.7% | 94.5% | 95.0% | 0.6% |
|  | D | 5 mos | 95% | 95.0% | 0.8% | 0.8% | 94.8% |  |  |
|  |  | 10 mos | 95% | 95.2% | 0.9% | 0.9% | 93.6% | 95.0% | 0.7% |
| (b) 5-mos $VE_a = 85\%$<br>10-mos $VE_a = 75\%$ | A | 5 mos | 85% | 84.9% | 1.4% | 1.5% | 95.4% | | |
|  |  | 10 mos | 75% | 74.9% | 2.2% | 2.2% | 94.9% | 78.5% | 1.4% |
|  | B | 5 mos | 85% | 85.0% | 1.3% | 1.3% | 95.1% |  |  |
|  |  | 10 mos | 75% | 75.0% | 2.6% | 2.6% | 94.9% | 81.6% | 1.3% |
|  | C | 5 mos | 85% | 85.0% | 1.3% | 1.3% | 95.7% |  |  |
|  |  | 10 mos | 75% | 75.0% | 2.4% | 2.4% | 94.8% | 80.9% | 1.3% |
|  | D | 5 mos | 85% | 85.0% | 1.5% | 1.5% | 94.7% |  |  |
|  |  | 10 mos | 75% | 74.9% | 3.4% | 3.4% | 94.6% | 84.6% | 1.4% |
| (c) 5-mos $VE_a = 70\%$<br>10-mos $VE_a = 50\%$ | A | 5 mos | 70% | 69.9% | 2.2% | 2.2% | 95.1% | | |
|  |  | 10 mos | 50% | 49.9% | 3.4% | 3.3% | 94.5% | 57.2% | 2.2% |
|  | B | 5 mos | 70% | 69.9% | 2.0% | 2.0% | 95.0% |  |  |
|  |  | 10 mos | 50% | 50.0% | 4.1% | 4.1% | 95.5% | 64.0% | 2.0% |
|  | C | 5 mos | 70% | 70.0% | 2.0% | 2.0% | 95.0% |  |  |
|  |  | 10 mos | 50% | 49.9% | 3.8% | 3.8% | 94.7% | 62.5% | 2.0% |
|  | D | 5 mos | 70% | 70.0% | 2.3% | 2.3% | 94.8% |  |  |
|  |  | 10 mos | 50% | 49.9% | 5.1% | 5.1% | 94.9% | 69.5% | 2.2% |

Note: Mean and SE denote the mean and standard error of the estimator for $VE_a$ , SEE denotes the mean of the standard error estimator, and CP denotes the coverage probability of the 95% confidence interval.

**Figure 2.**
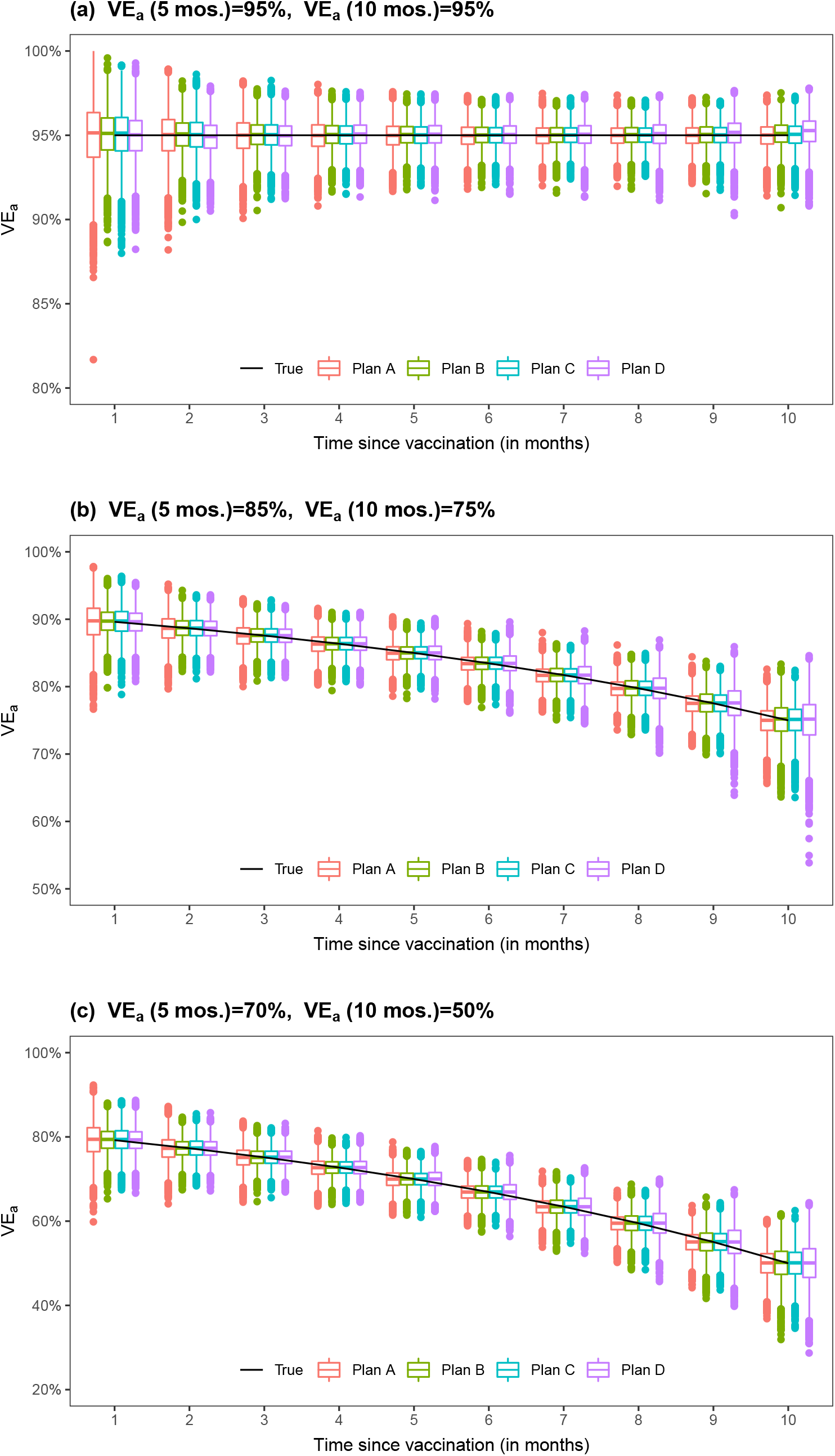
Proposed estimates of VE_a_ under no crossover and three blinded crossover plans.

We also evaluated the performance of standard Cox regression,^17−18^ using vaccine status as a potentially time-dependent covariate with a constant hazard ratio. Because it estimates an overall VE among individuals who have been vaccinated for different amounts of time, this method over-estimates long-term VE when VE decreases over time, although the estimation is unbiased when VE is constant over time. Even though the results for standard Cox regression are shown in the “10-mos.” row of the table, the estimates do not pertain to the hazard ratio at month 10 or to the average hazard ratio over the first 10 months. In scenario (c), where 10-month VE_a_ equals 50% and month-10 VE_h_ equals –3.8%, standard Cox regression yields mean VE estimates of 57.2%, 64.0%, 62.5%, and 69.5% under Plans A, B, C, and D, respectively; the poor performance under Plan A highlights the fact that standard Cox regression does not properly capture waning VE even when all placebo recipients remain on their original assignments until the end of the trial.

We conducted a second set of simulation studies by considering four unblinded crossover designs:

B’. Crossover occurs at month 6.5, 7.5, 8.5, 9.5, or 10.5 for participants with priority tier of 1, 2, 3, 4, or 5, respectively.

C’. 20% of participants follow Plan A, and the rest follow Plan B’. D’. Crossover occurs at month 6.5 for all participants.

D”. Crossover starts at month 6 for all participants, with the waiting time following the exponential distribution with mean of 0.5 month.

With unblinded crossover, participants are notified of their original treatment assignments at the point of crossover, and placebo recipients are vaccinated soon after. In Plan B’, everyone in a given priority-tier group is notified of their original treatment assignment on the same day. In Plan D’, all participants are notified of their original treatment assignments on the same day. In Plan D”, participants are notified of their randomization assignments without priority tiering; the timing of crossover is the same as that of Plan D. Plan D” was meant to mimic the crossover that has been occurring in the two mRNA vaccine trials, where participants have been unblinded gradually.

Because vaccine recipients may engage in riskier behavior upon unblinding and placebo recipients may also change their behavior upon unblinding (in a manner that likely differs from vaccine recipients), we discarded the data collected after unblinding for both the vaccine and placebo groups by censoring each participant’s time to disease at their time of unblinding. This strategy avoids any bias due to behavioral confounding.

The results for the estimation of VE_a_ based on 10,000 simulated datasets are summarized in Table 2 and Figure 3. The proposed method yields unbiased estimates of the VE_a_ curves under Plans B’ and C’, but the standard errors for estimates of long-term VE are markedly higher than under Plans B and C; the standard errors estimates are accurate only up to month 8. Under Plan D’, where all placebo recipients are vaccinated at month 6.5, the estimates of 5-month VE_a_ are unbiased; however, the estimates of 8-month or 10-month VE_a_ are biased, especially when VE wanes over time. (Using the unblinded data would not reduce the bias in estimating long-term VE because there are no placebo participants beyond month 6.5.) Of note, the bias of standard Cox regression in estimating waning VE is more severe under unblinded crossover than under blinded crossover. This result reflects the fact that standard statistical methods do not provide valid estimates of waning VE even in the absence of crossover — because time to disease is censored at time of unblinding, there is no actual crossover of treatments in this analysis.

**Table 2.**
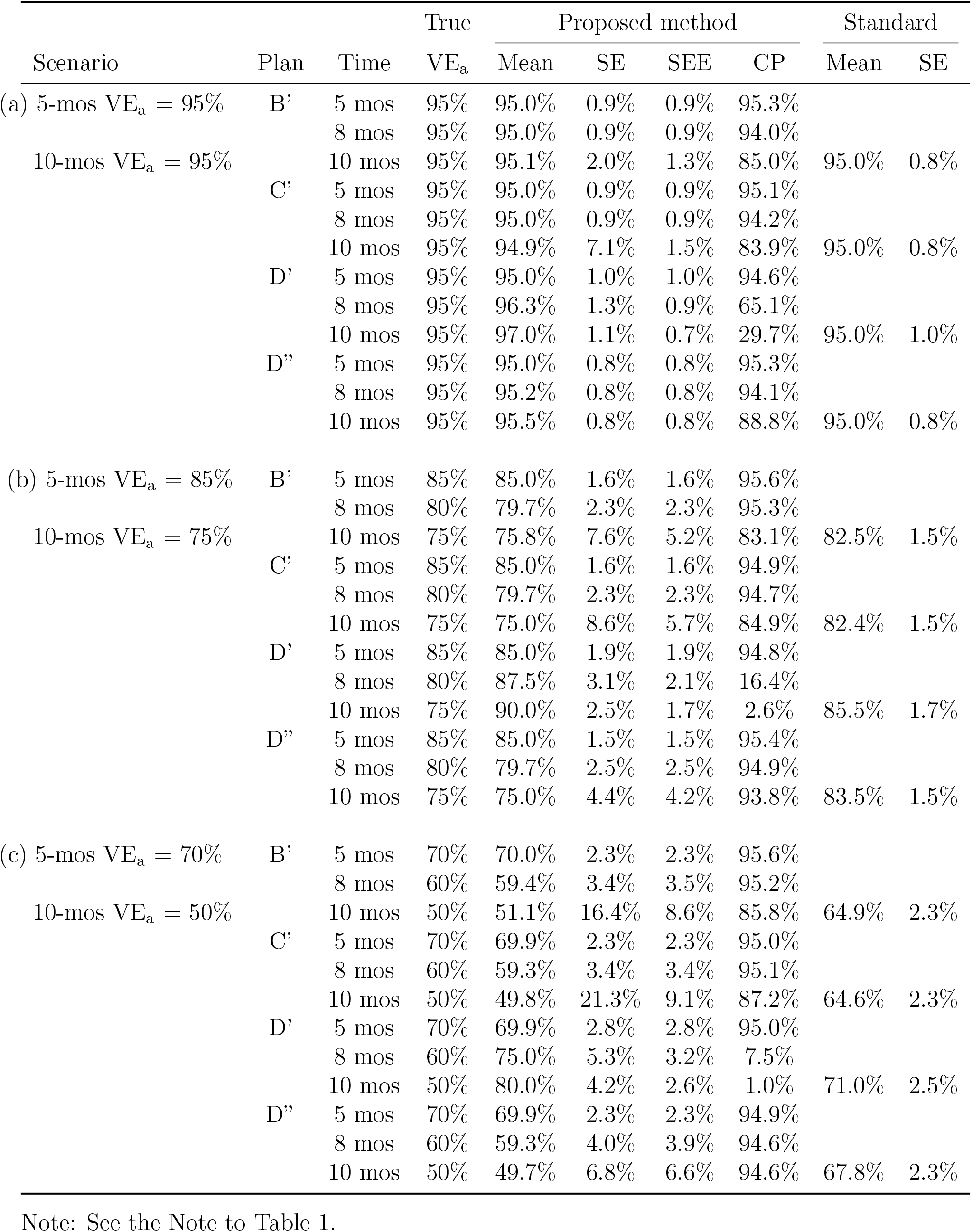
**Estimation of Time-Varying** VE_a_ **by the Proposed and Standard Methods Under Four Unblinded Crossover Plans**

**Figure 3.**
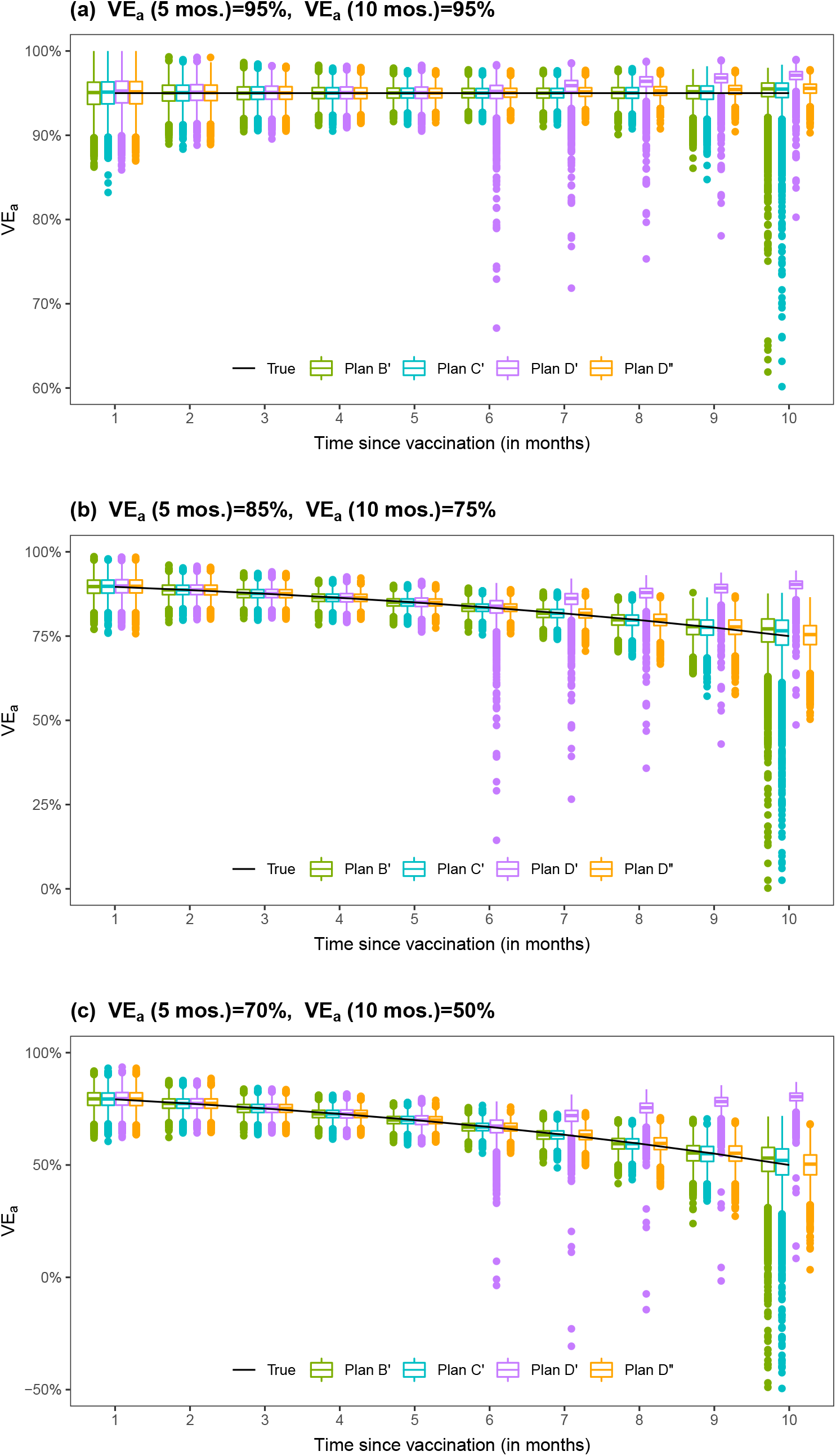
Proposed estimates of VE_a_ under four unblinded crossover plans.

The results under Plan D” are encouraging: the estimates of the VE_a_ curves have little bias, and the standard errors are accurately estimated, such that confidence intervals have proper coverage probabilities — at least for the first 8 months. The timing of crossover is the same between Plans D and D”; however, we disregarded the follow-up data collected after unblinding, such that the precision of estimates is lower under Plan D” than under Plan D. Although the mean time to crossover under Plan D” is the same as that of Plan D’, crossover spreads over a longer period under Plan D”, making it possible to estimate long-term VE.

Figures 4 and 5 show the results for estimating VE_h_ from the first and second sets of simulation studies, respectively. Under Plans A–D, the estimates of the VE_h_ curves are virtually unbiased, at least up to month 9. Under Plans B’ and C’, the VE_h_ estimates are nearly unbiased up to month 8. Under Plan D’, VE_h_ is estimated well until month 5. Under Plan D”, the estimates of the VE_h_ curves are virtually unbiased, except for the last 2 months in scenario (a).

**Figure 4.**
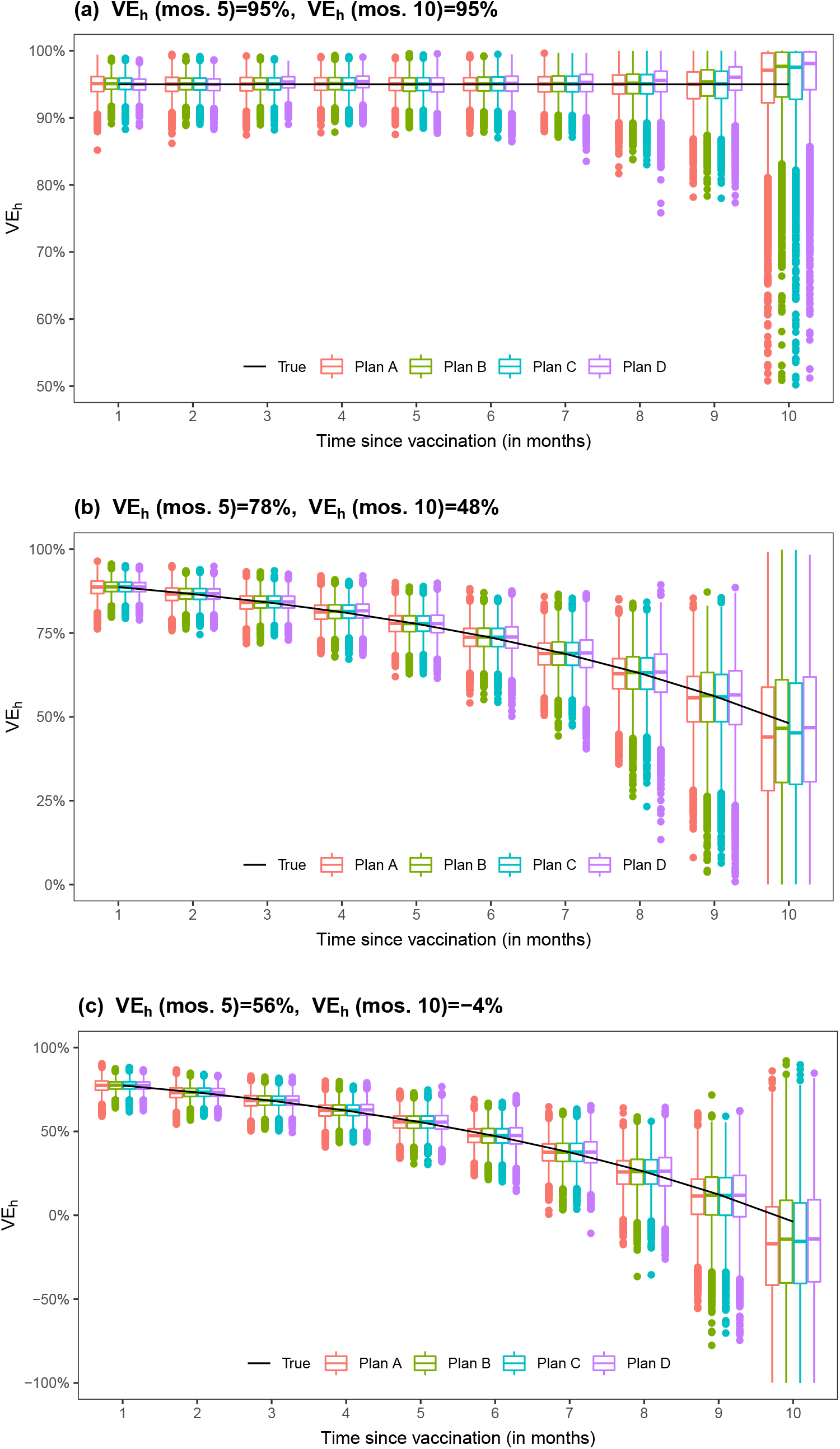
Proposed estimates of VE_h_ under no crossover and three blinded crossover plans.

**Figure 5.**
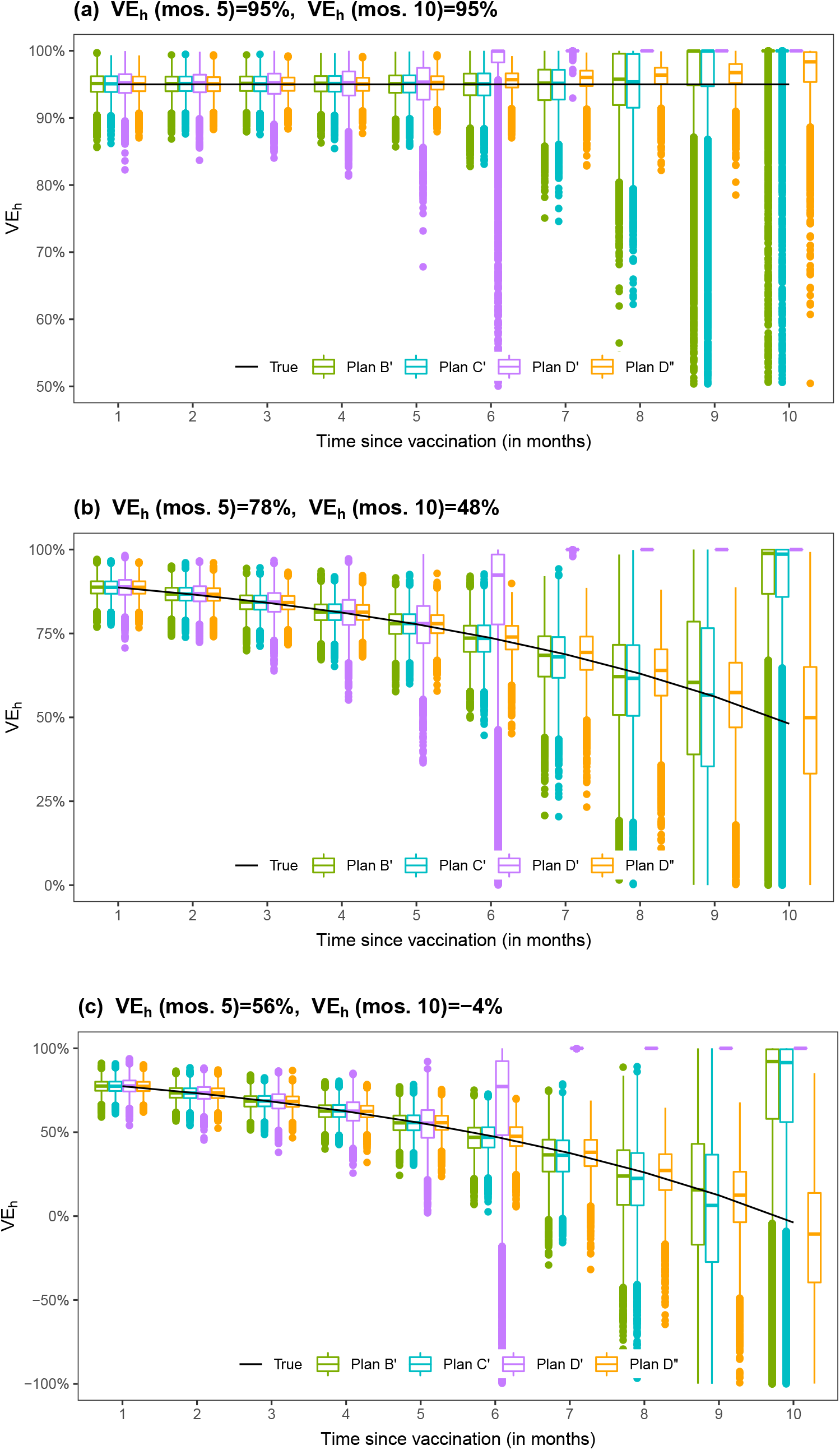
Proposed estimates of VE_h_ under four unblinded crossover plans.

Figure 6 displays the estimation results for VE_a_ produced by the proposed methods in one of the trials simulated with 75% 10-month VE_a_. The estimates of VE_a_ are close to the truth, and the 95% confidence intervals cover the truth up to the end of crossover. In terms of estimating long-term VE_a_, Plan C is nearly as good as Plan A and is slightly better than Plan B, which is better than Plan D; Plans B, C, and D are considerably better than Plans B’, C’, and D’, respectively; and Plan D” is much better than Plan D’.

**Figure 6.**
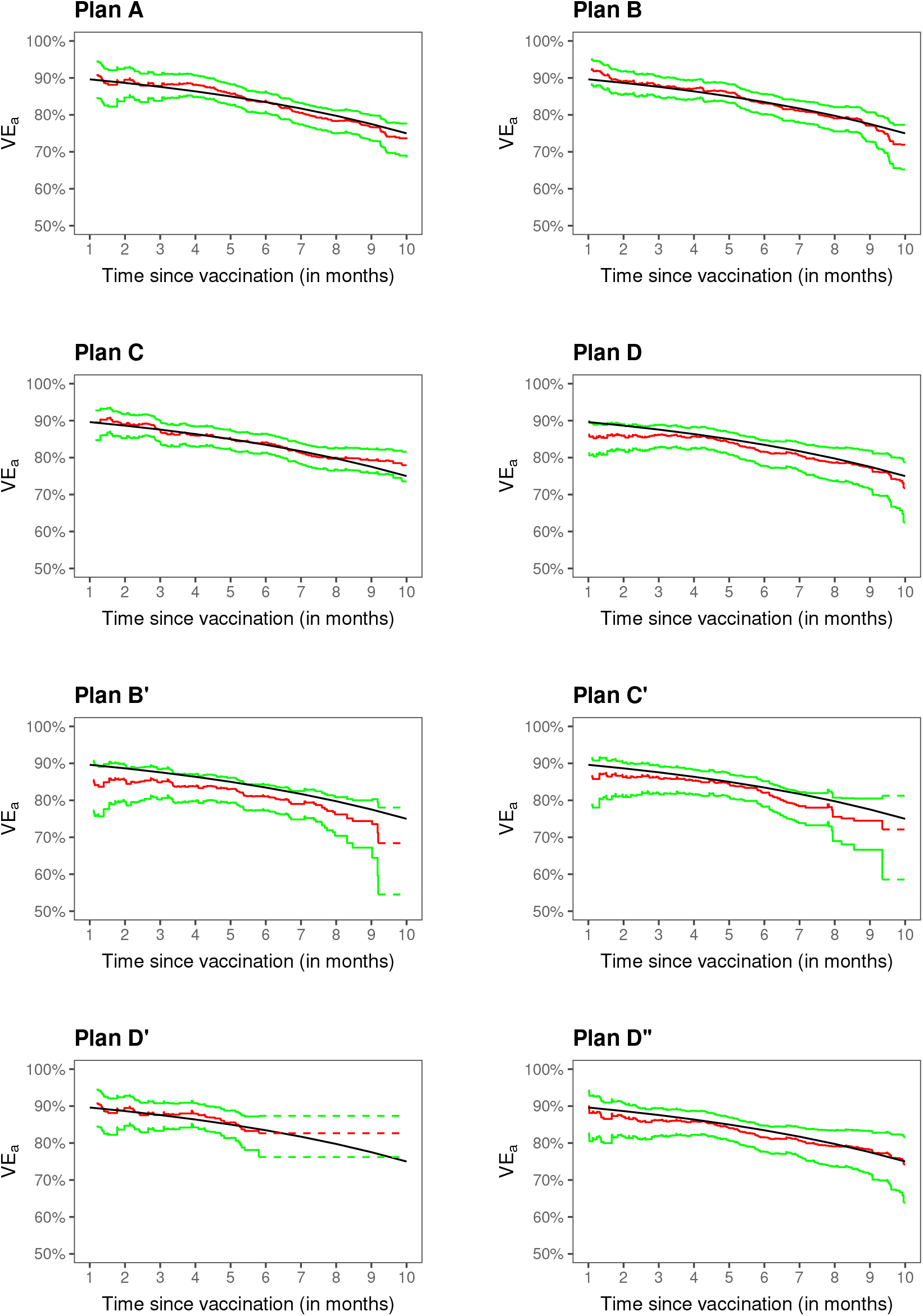
Estimation of VE_a_ a clinical trial with no crossover (A), three blinded crossover (B–D), and four unblinded crossover (B’–D”): the black curve pertains to the true value, the red curve to the proposed estimate, and the green curves to the 95% confidence intervals.

Figure 7 presents the analysis results for VE_h_ from the same trial. The estimates are close to the truth up to the point where there are still a few placebo participants under follow-up. The estimates are more bumpy under unblinded than blinded crossover. Comparing Figures 6 and 7 shows that VE_h_ is much lower than VE_a_ in the presence of waning vaccine effect.

**Figure 7.**
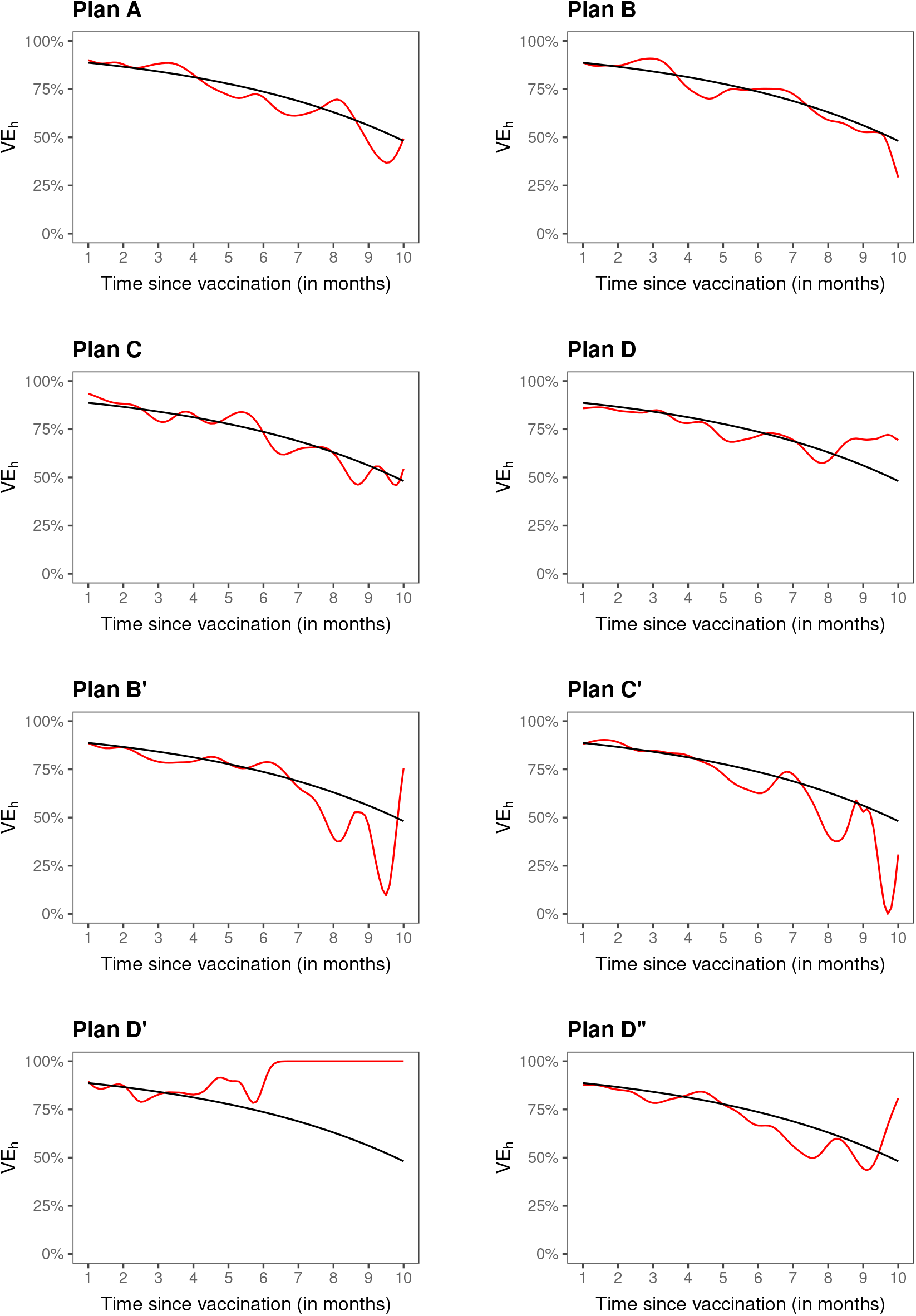
Estimation of VE_h_ a clinical trial with no crossover (A), three blinded crossover (B–D), and four unblinded crossover (B’–D”): the black curve pertains to the true value, and the red curve to the proposed estimate.

## Discussion

For a preventive COVID-19 vaccine to be administered to millions of people, including healthy individuals, its safety and efficacy must be demonstrated in a clear and compelling manner. Although preliminary results from ongoing phase 3 clinical trials have revealed higher than expected efficacy of COVID-19 vaccines,^4–6^ additional follow-up is required to assess long-term efficacy and safety. Indeed, FDA does not consider issuance of an EUA, in and of itself, as grounds for stopping blinded follow-up in an ongoing clinical trial.^15^

We recommend the rolling crossover design, which allows placebo volunteers to be vaccinated in a timely manner while still making it possible to assess long-term vaccine safety and efficacy. As our simulation studies have shown, standard Cox regression with a constant hazard ratio seriously over-estimates long-term VE in the presence of waning vaccine effect. We have developed a valid and efficient approach to evaluate the effect of a COVID-19 vaccine that potentially wanes over time. The estimated curve of time-varying VE can be used to determine when a booster vaccination is needed to sustain protection; this information is also an important input parameter in mathematical modeling of the population impact of COVID-19 vaccines.

We have considered vaccine effects on cumulative incidence and on instantaneous risk. It is also of interest to study vaccine effects over successive time periods. We present the method and results for estimating period-specific VE in Supplemental Appendices 1 and 2.

To ensure high-quality follow-up data, crossover should ideally be blinded, with participants not knowing their treatment assignments even after crossover. It is advantageous, when possible, to implement crossover on a rolling basis rather than instantaneously since time-varying VE can be estimated (without adding assumptions) only up to the point where there are still a few placebo recipients under follow-up. Indeed, rolling crossover is even more important than blinding for the express purpose of assessing long-term VE without imposing additional assumptions.

Of course, unblinded crossover has practical benefits over blinded crossover: it reduces operational complexity and trial cost. However, unblinding can lead to differential exposure to SARS-CoV-2 between the original vaccinees and the placebo crossovers, which in turn can bias the estimation of VE. This bias can be avoided by analyzing only the blinded follow-up data. However, discarding the unblinded follow-up data may substantially reduce the precision in estimating long-term VE. We may estimate VE twice, once with all follow-up data and once with only blinded follow-up data, and compare the two sets of results.

Alternatively, our methods can be applied to all follow-up data, followed by a sensitivity analysis to assess the robustness of the results to potential unmeasured confounding caused by unblinding of trial participants. In Supplemental Appendix 3, we show how to apply a best-practice general methodology in epidemiological research^19−20^ to perform this sensitivity analysis. Using this methodology, we can assess how strong unmeasured confounding due to unblinding would need to be in order to fully explain away the observed VE. We can also provide a conservative estimate of VE that accounts for unmeasured confounding.

Recently, Follmann et al ^21^ advocated blinded crossover and continued follow-up of trial participants to assess vaccine durability and potential delayed enhancement of disease. They estimated an overall VE for the original vaccine arm in the post-crossover period (after all placebo volunteers were vaccinated) by imputing the case count for a counter-factual placebo group under certain assumptions. We address the issue of VE durability in a different way, using observed data to estimate the entire curve of VE as a function of time elapsed since vaccination, up to the point where most participants have been vaccinated. Our approach requires minimal assumptions and is applicable to both blinded and unblinded crossover plans, with any length of additional follow-up.

The methods we have described assess overall VE against all viral variants of COVID-19. The results may be difficult to interpret if the distribution of viral variants changes during the trial, because waning VE might be caused by increasing prevalence of resistant variants while VE against specific variants remains constant. It is possible to address this challenge if SARS-CoV-2 sequences from all COVID-19 cases are measured. Our methods can then be applied to the subset of COVID-19 cases caused by each specific variant or set of variants. Conducting this analysis for each spectrum of viral genotypes provides interpretable results on the durability of VE.

Although we have focused on VE in the entire study population, we can also estimate VE for various subgroups, such as age group, sex, and race/ethnic group, by applying our methods to a subset of participants. Because we allow the effect of vaccination to vary over time in an arbitrary manner, however, our estimates of long-term VE may be unstable if there are only a small number of cases in a subgroup. To alleviate this problem, we may formulate the time-varying hazard ratio through a parametric (e.g., log-linear) function, which is then allowed to interact with subgroups.

We have targeted VE over the first 10 months for several reasons. First, it is unlikely that there will be any placebo volunteers beyond month 10 in the ongoing COVID-19 vaccine trials, and follow-up data in the absence of a placebo arm do not provide direct information about VE durability. Second, estimates of long-term VE will become more uncertain as community transmission decreases.

Although we have framed the discussion in the context of randomized, placebo-controlled phase 3 trials, our methods do not require the designation of vaccine and placebo groups; instead, everyone is considered a potential vaccinee, with the only distinction being *when* they are vaccinated. This general framework can be applied to surveillance data in order to estimate the risk of disease as a function of time elapsed since vaccination in the “real world”. With the large volume of surveillance data, we can estimate the effectiveness of vaccination in various sub-populations and against different viral variants, as well as the duration of any protective effect. Causal interpretations of results, however, require the timing of vaccination to depend only on observed covariates. If it is impossible to measure the key timing factors or inappropriate to include them as covariates in the model, then the aforementioned sensitivity analysis would be warranted.

We have implemented the methods described in this paper in an R package, which is available at https://dlin.web.unc.edu/software/dove/.

## Data Availability

N/A

https://dlin.web.unc.edu/software/dove/

## Acknowledgments

The authors are grateful to Yu Gu and Bridget I. Lin for assistance, to Thomas Fleming, David Harrington, and Ross Prentice for helpful discussions, and to the editors and two referees for fast reviews. This work was supported by the National Institutes of Health grants R01 AI029168, R01 GM124104, P01 CA142538, and UM1 AI068635.

## Supplemental Appendix

### 1. Vaccine Efficacy

Let *S* denote the time when vaccination takes place and *T* denote the time when symptomatic COVID-19 develops; both times are measured in days from the start of the clinical trial (Figure 1). (We measure each participant’s time to disease from the start of the trial rather than from their time of enrollment because the risk of disease depends on community transmission, which varies over the calendar time. Obviously, the two time scales are different under staggered enrollment.) In addition, let *X* denote baseline risk factors (e.g., age, occupation, comorbidities). We specify that the hazard function of *T* is related to *S* and *X* through the Cox regression model [1] with a time-varying vaccine effect

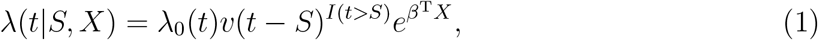

where *λ*_0_(*·*) is an arbitrary baseline hazard function, *v*(*·*) is a positive function characterizing the time-varying effect of vaccination, *β* is a set of regression parameters representing the effects of baseline risk factors, and *I*(*·*) is the indicator function. Under this formulation, the baseline hazard rate varies over the calendar time; the effect of vaccine on the risk of disease depends on the time elapsed since vaccination, but not on the specific date of vaccination or on the baseline risk factors. (The latter restriction can be relaxed by applying the model separately to each sub-population of interest.)

Vaccine efficacy (VE) at time *t* after vaccination is generally defined by *V E*(*t*) = 1 *− RR*(*t*), where *RR*(*t*) is some measure of relative risk at time *t* comparing the vaccinated population to the unvaccinated population [2,3]. The most common measures of risk in vaccine trials are hazard rate and attack rate [3,4]. In our formulation, the relative hazard rate or hazard ratio at time *t* is simply *v*(*t*). The relative attack rate at time *t* is the ratio of the cumulative incidence of disease at time *t* for individuals who have been vaccinated for *t* days compared with those who are unvaccinated:

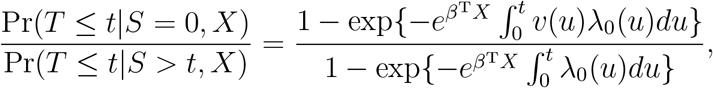

which is approximately

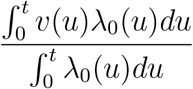

when the incidence is low. (This condition holds for Covid-19 vaccine trials because the annualized incidence of symptomatic COVID-19 is less than 5%.) If *λ*_0_(*·*) is approximately constant, then the above ratio can be written as 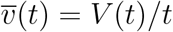, where

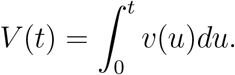

Clearly, 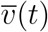 is the average hazard ratio over the time period (0, *t*].

Thus, we have two definitions of time-varying VE: one in terms of hazard rate

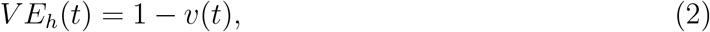

and one in terms of attack rate

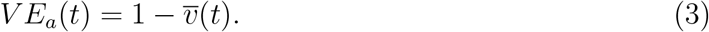

Note that the assumption of an approximately constant baseline hazard rate over time is needed in order to interpret 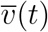 as the ratio of the cumulative incidence but is not used in estimation. We will refer to *V E*_*a*_(*t*) as the VE in attack rate whether the baseline hazard rate is constant or not. We call *V E*_*h*_(*t*) the day-*t* VE, which pertains to the instantaneous vaccine effect at time *t*, and call *V E*_*a*_(*t*) the *t*-day VE, which pertains to the cumulative vaccine effect over the time interval (0, *t*]. If the hazard ratio is constant over time, then the two definitions are equivalent. If the hazard ratio increases over time, then *V E*_*a*_(*t*) will be higher than *V E*_*h*_(*t*).

Suppose that the clinical trial enrolls a total of *n* participants. For *i* = 1,…, *n*, let *R*_*i*_, *T*_*i*_, *S*_*i*_, *C*_*i*_, and *X*_*i*_ denote, respectively, the entry time, the time to symptomatic COVID-19, the time to vaccination, the time to loss to follow-up, and the baseline risk factors for the *i*th participant. The data consist of (*R*_*i*_, *Y*_*i*_, Δ_*i*_, *D*_*i*_, *D*_*i*_*S*_*i*_, *X*_*i*_) (*i* = 1,…, *n*), where *Y*_*i*_ = min(*T*_*i*_, *C*_*i*_), Δ_*i*_ = *I*(*T*_*i*_ ≤ *C*_*i*_), and *D*_*i*_ = *I*(*S*_*i*_ ≤ *Y*_*i*_).

We assume that (*R*_*i*_, *S*_*i*_, *C*_*i*_) are independent of *T*_*i*_ conditional on *X*_*i*_. The likelihood takes

the form

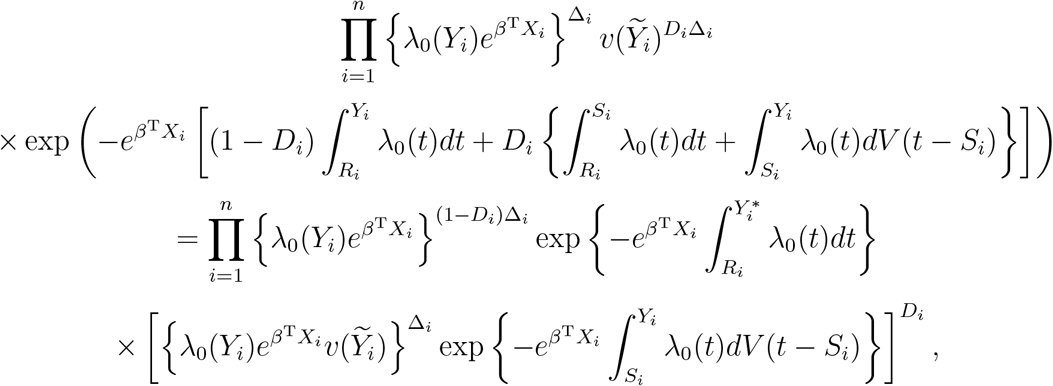

where 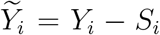, and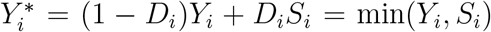. We approximate log *λ*_0_(*t*) through splines with *m* basis functions, *B*_1_(*t*),…, *B*_*m*_(*t*), such that 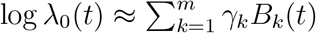 Let *θ* = (*β*^T^, *γ*_1_,…, *γ*_*m*_)^T^, and 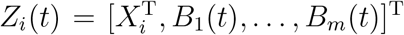. We perform the nonparametric maximum likelihood estimation [5], in which *V* (*·*) is treated as a step function jumping at the time points 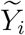 with *D*_*i*_ = Δ_*i*_ = 1. Thus, we maximize the objective function

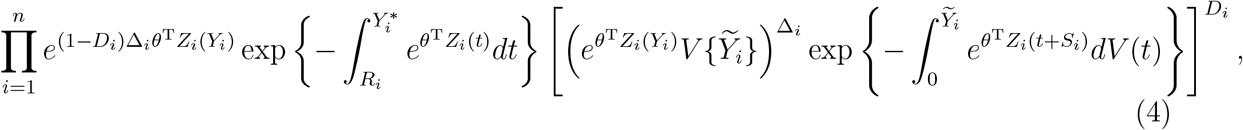

where *V* {*t*} is the jump size of *V* (*·*) at *t*.

We first maximize the objective function in (4) for fixed *θ* to yield

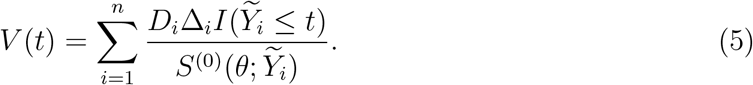

Here and in the sequel, 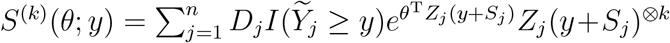, where *a*^⊗0^ = 1, *a*^⊗1^ = *a*, and *a*^⊗2^ = *aa*^T^ for a column vector *a*. After plugging (5) into (4), we obtain the profile likelihood for *θ*. Differentiating the profile log-likelihood with respect to *θ* yields the estimating function

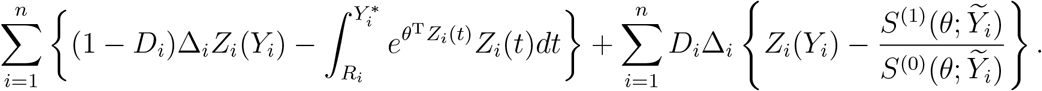

Denote the resulting estimator of *θ* by 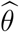. Replacing *θ* in (5) by 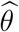 yields the estimator of *V* (*t*)

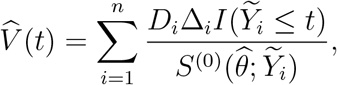

which is reminiscent of the Breslow estimator [6] for the cumulative baseline hazard function under the standard Cox model. We then estimate *V E*_*a*_(*t*) through equation (3).

Using counting-process martingale theory and other mathematical arguments [7,8], we can show that 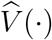is consistent and asymptotically normal. In addition, the covariance between 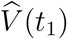 and 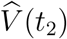 can be consistently estimated by 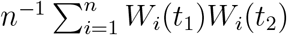, where

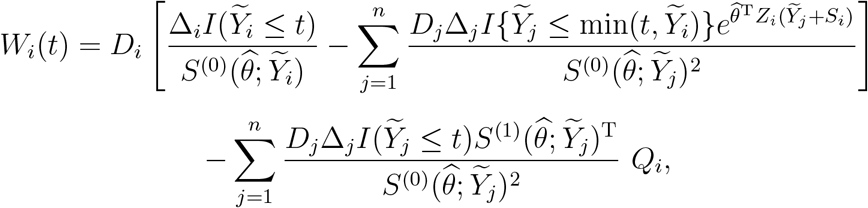

and

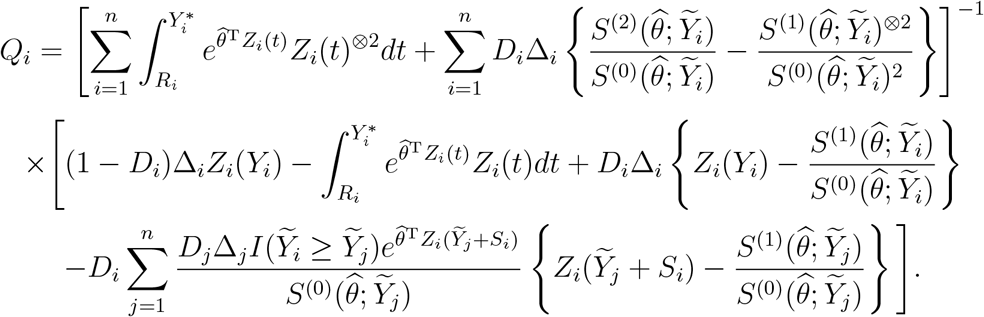

These results can be used to construct confidence intervals for *V E*_*a*_(*t*). Specifically, we first construct the confidence intervals for log{*V* (*t*)} and then transform them to *V E*_*a*_(*t*). We can also construct simultaneous confidence bands for *V E*_*a*_(*·*) [8].

We estimate *v*(*·*) by applying a local linear smoother to the jump sizes of 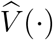. The jump size of 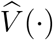 at 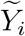_*i*_ is given by 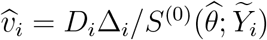. For any *t*, we fit a local linear regression model and estimate its intercept *A*(*t*) and slope *B*(*t*) by solving the equation

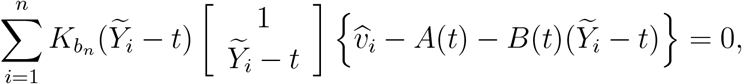

where 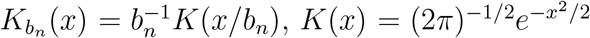, and *b*_n_ is a data-adaptive bandwidth. We let 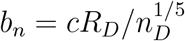, where *c* is a tuning parameter, *R*_*D*_ is the range of the observed event times, and *n*_*D*_ is the number of disease cases [9]. Denote the resulting estimator of *A*(*t*) by *Â* (*t*). We then estimate *v*(*t*) by

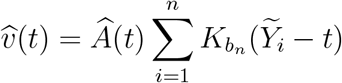

and estimate VE_h_(t) according to equation (2). We let the tuning parameter *c* lie between 0.1 and 1.0. A smaller value of *c* makes 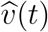 less biased but more variable, whereas a larger value of *c* generates a more smooth estimate of the VE_h_ curve.

Finally, we define the VE in attack rate over the time interval (*t*_1_, *t*_2_] by

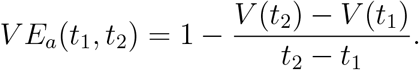

The fraction on the right-hand side is the average hazard ratio over the time period (*t*_1_, *t*_2_]. We estimate 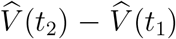, which is normally distributed with variance 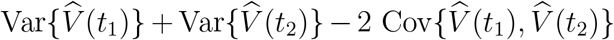. We then construct the confidence intervals for *V E*_*a*_(*t*_1_, *t*_2_) based on the log-transformation.

*Remark*. Durham et al. [2] considered a special case of model (1) in which all participants are vaccinated at approximately the same time (such that *S* can be set 0 for all participants) and estimated the time-varying hazard ratio by smoothing the residuals from the standard Cox model with a constant hazard ratio. If community transmission is constant over time, then one can estimate the incidence rate in the vaccine group by using the number of cases among the participants who have been vaccinated for the same amount of time and assess waning VE by comparing the estimates of relative incidence rates for successive time periods. In COVID-19 vaccine trials, the enrollment period is relatively long compared with the study duration, and community transmission varies considerably over time. Thus, it is necessary to adopt two different time scales: time since study initiation for the disease endpoint and time since vaccination for the vaccine effect.

## 2. Simulation Studies

We assumed that 40,000 participants entered the study at a constant rate over four months, i.e., *R ∼* Uniform(0, 4). We created a composite baseline risk score *X*, which takes values 1, 2, 3, 4, and 5 with equal probability. At study entry, half of the participants were assigned to vaccine and half to placebo. The statistically optimal design would be to maintain the original vaccine and placebo groups until the end of the study; we refer to this design as Plan A. We also considered three blinded crossover designs, under which placebo participants receive vaccine and vaccine participants receive placebo at the time of crossover, and all participants are followed until the end of the study or the time of analysis, which was set to be 10.5 months. The three blinded crossover designs are as follows:

Plan B. Crossover occurs at month (11 *− X* + *G*), where *G* follows the exponential distribution with mean of 0.5 month.

Plan C. 20% of participants follow Plan A and 80% follow Plan B.

Plan D. Crossover occurs at month 6 + *G*, where *G* follows the exponential distribution with mean of 0.5 month.

Plan C mimics a scenario where all participants are offered the option of crossover, but a small percentage (20%) choose to stay on the original assignments.

In addition, we considered four unblinded crossover designs:

Plan B’. Crossover occurs at month (11.5 *− X*).

Plan C’. 20% of participants follow Plan A and 80% follow Plan B’.

Plan D’. Crossover occurs at month 6.5.

Plan D”. Crossover occurs at the same time as Plan D.

Under unblinded crossover, participants are notified of their original assignments at the time of crossover, and placebo participants receive the vaccine soon after. In practice, only placebo participants would cross over, since there is no need to give placebo to vaccine recipients after unblinding. In Plans B’–D”, the time of crossover is the time of unblinding rather than the time when placebo participants actually receive the vaccine. Because participants might change their behavior upon discovering their original treatment assignments, we discarded the follow-up data collected after unblinding by censoring each participant’s time to disease at their time of unblinding. This strategy avoids bias due to behavioral confounding at the cost of reduced statistical efficiency.

We generated the event time *T* from m

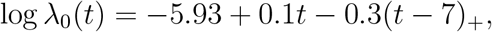

And

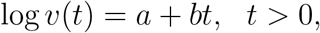

where *t*_+_ = *t* if *t >* 0 and 0 otherwise, and *a* and *b* were chosen to achieve the desired 5-month and 10-month VE_a_. We censored *T* at the time of unblinding under Plans B’–D”.

For each simulated dataset, we estimated log *λ*_0_(*t*) using a piece-wise constant function with 20 pieces placed at the equal quantiles of the observed event times. We then estimated *V* (*t*) using the proposed method and estimated *V E*_*a*_(*t*) according to equation (3). In addition, we estimated *v*(*t*) through local linear regression with tuning parameter *c* = 0.1 and estimated *V E*_*h*_(*t*) according to equation (2). For comparison, we also fit the standard Cox proportional hazards model that includes *X* and time-dependent covariate *I*(*S < t*) with a constant hazard ratio and estimated VE by 1 minus the estimated hazard ratio of the time-dependent covariate. The results are reported in the main text.

We also evaluated our method for estimating *V E*_*a*_(*t*_1_, *t*_2_) over a partition of the study period in the simulation studies. We found that the method performs well when the time interval is at least one-month wide. For example, Table S.1 summarizes the results for estimating monthly *V E*_*a*_ under Plan D” in scenario (b). The *V E*_*a*_ estimates are virtually unbiased, the standard errors are accurately estimated, and the confidence intervals have proper coverage probabilities. Table S.2 shows the analysis results for bimonthly *V E*_*a*_ in one simulated trial. Bimonthly *V E*_*a*_ is estimated with higher precision than monthly *V E*_*a*_.

## 3. Sensitivity Analysis

We suggest reporting the E-value [10] as a summary measure of the evidence against the null hypothesis *H*_0_: *V E*(*t*) ≤ 0. The E-value is the minimum strength of association on the risk ratio scale, i.e, *RR*(*t*) = 1 *− V E*(*t*), that an unmeasured confounder would need to have with both vaccination status and disease outcome in order to fully explain away a specific observed VE. Let 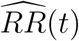) be the estimate of *RR*(*t*), and let *UL*(*t*) be the upper limit of the 95% confidence interval for *RR*(*t*). Then the E-value for 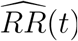 is given by

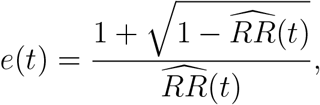

provided that 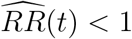. In addition, the E-value for *UL*(*t*) is computed as 1 if *UL*(*t*) ≥ 1 and as

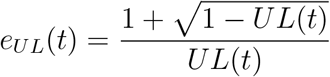

otherwise. E-values near one suggest weak support for a causal inference, and greater E-values provide increasing evidence for causality.

Suppose, for example, that 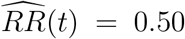, with 95% confidence interval (0.08, 0.75). Then *e*(*t*) = 3.4, meaning that the result of 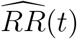 being less than one could be explained away by an unmeasured confounder associated with both vaccination status and disease by a risk ratio of 3.4-fold each after accounting for the vector *X* of measured confounders, but not by a weaker unmeasured confounder. In addition, *e*_*UL*_(*t*) = 2, which is the strength of unmeasured confounding at which statistical significance for *V E*(*t*) *>* 0 would be lost. These two E-values judge how confident we can be that *V E*(*t*) truly exceeds 0, accounting for potential unmeasured confounding due to unblinding and for sampling variability.

We can provide a conservative estimate of *V E*(*t*) that accounts for potential unmeasured confounding [11]. Let *RR*_*UD*_(*t*) be the maximum risk ratio for disease when comparing any two categories of the unmeasured confounder *U*, within either the vaccinated group or the unvaccinated group, conditional on the vector *X* of observed covariates, and let *RR*_*EU*_ (*t*) be the maximum risk ratio for any specific level of the unmeasured confounder *U* when comparing the vaccinated and unvaccinated individuals. Of note, *RR*_*UD*_(*t*) quantifies the importance of the unmeasured confounder *U* for disease, and *RR*_*EU*_ (*t*) quantifies how imbalanced the vaccinated and unvaccinated groups are in the unmeasured confounder *U*. We define the bias or bounding factor

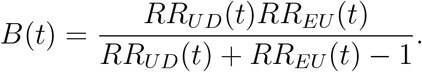

Then a conservative (lower bound) estimate of the VE is given by 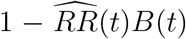, and a conservative confidence interval is obtained by multiplying the lower and upper limits of the confidence interval for *RR*(*t*) by *B*(*t*).

---

**Table S1.** Estimation of Monthly VE_a_ Under Unblinded Crossover Plan D”.

| Months | True $VE_a$ | Bias | SE | SEE | CP |
| --- | --- | --- | --- | --- | --- |
| 0–1 | 89.6% | 0.0% | 2.9% | 2.9% | 95.4% |
| 1–2 | 87.7% | -0.1% | 3.0% | 3.0% | 95.6% |
| 2–3 | 85.4% | 0.0% | 3.0% | 3.1% | 95.8% |
| 3–4 | 82.7% | -0.1% | 3.3% | 3.3% | 95.3% |
| 4–5 | 79.5% | -0.1% | 3.8% | 3.8% | 95.3% |
| 5–6 | 75.8% | -0.2% | 4.8% | 4.8% | 95.5% |
| 6–7 | 71.3% | -0.1% | 6.6% | 6.5% | 95.1% |
| 7–8 | 66.0% | 0.0% | 9.2% | 9.0% | 95.0% |
| 8–9 | 59.7% | 0.1% | 12.8% | 12.6% | 95.2% |
| 9–10 | 52.3% | 0.3% | 19.4% | 18.9% | 95.2% |

Note: Bias and SE denote the bias and standard error of the estimator for $VE_a$ , SEE denotes the mean of the standard error estimator, and CP denotes the coverage probability of the 95% confidence interval.

**Table S2.** Estimation of Bimonthly VE_a_ in a Clinical Trial.

| Months | Estimate | Std Error | 95% Conf Interval |
| --- | --- | --- | --- |
| 0–2 | 88.7% | 2.1% | (83.7%, 92.2%) |
| 2–4 | 88.1% | 2.0% | (83.6%, 91.4%) |
| 4–6 | 80.4% | 2.9% | (74.0%, 85.3%) |
| 6–8 | 78.5% | 4.9% | (66.5%, 86.2%) |
| 8–10 | 67.4% | 9.5% | (42.2%, 81.6%) |

